# Bayesian Spatio-temporal prediction and counterfactual generation: an application in non-pharmaceutical interventions in Covid-19

**DOI:** 10.1101/2022.11.30.22282938

**Authors:** Andrew Lawson, Chawarat Rotejanaprasert

## Abstract

The spatio-temporal course of an epidemic (such as Covid-19) can be significantly affected by non-pharmaceutical interventions (NPIs), such as full or partial lockdowns. Bayesian Susceptible-Infected-Removed (SIR) models can be applied to the spatio-temporal spread of infectious disease (STIF) (such as Covid-19). In causal inference it is classically of interest to investigate counterfactuals. In the context of STIF it is possible to use nowcasting to assess the possible counterfactual realization of disease in incidence that would have been evidenced with no NPI. Classic lagged dependency spatio-temporal IF models will be discussed and the importance of the ST component in nowcasting will be assessed. The real example of lockdowns for Covid-19 in two US states during 2020 and 2021 is provided. The degeneracy in prediction in longer time periods is highlighted and the wide confidence intervals characterize the forecasts.

## 1) Introduction

During the Covid-19 pandemic period of 2020 many countries worldwide enacted lockdowns to try to control the spread of the virus. These lockdowns are examples of non-pharmaceutical interventions (NPIs) and were used mainly prior to the availability of vaccination.

The CDC notes that:

> **Nonpharmaceutical Interventions (NPIs)** are actions, apart from getting vaccinated and taking medicine, that people and communities can take to help slow the spread of illnesses like pandemic influenza (flu). NPIs are also known as community mitigation strategies. When a new flu virus spreads among people, causing illness worldwide, it is called pandemic flu. Because a pandemic flu virus is new, the human population has little or no immunity against it. This allows the virus to spread quickly from person to person worldwide. NPIs are among the best ways of controlling pandemic flu when vaccines are not yet available. https://www.cdc.gov/nonpharmaceutical-interventions/

NPIs can take various forms and can extend for different time periods. In the US many southern states enacted lockdowns for only a few weeks, whereas northern states lockdowns were longer. In some cases only partial lockdowns were observed, whereby some businesses remained open but e. g. schools were shut. In the US state of South Carolina (SC) initial case reports in early March 2020, followed by the pandemic declaration by WHO (March 12^th^), led to state of emergency declaration on March 13^th^ and school closures on March 15^th^, restaurants closed on March 17^th^ and on March 19^th^ non-essential state employees and colleges to shelter in place.

Not until April 1^st^ did the state authorize closure of non -essential businesses. April 3^rd^ saw the introduction of travel restrictions, and on April 7^th^ a full lockdown with non-essential travel banned and work at home ordered.

By April 21^st^ retail stores were allowed to reopen and by May 4^th^ the home and work order was lifted and outdoor dining allowed. Finally, by June 11^th^ all restrictions lifted. In effect the main full lockdown lasted only 2 weeks.

Figure 1 displays the case count time profiles for Charleston and Richland counties in SC during the first part of the pandemic, for 353 days up to end of February 2021. Listed are early dates related to lockdowns in 2020. It is notable that following the full lifting of lockdowns in June 2020 there are significant increases in case counts leading into the large summer wave. Whether the partial or full lockdowns were effective in controlling early spread is difficult to ascertain.

**Figure 1.**
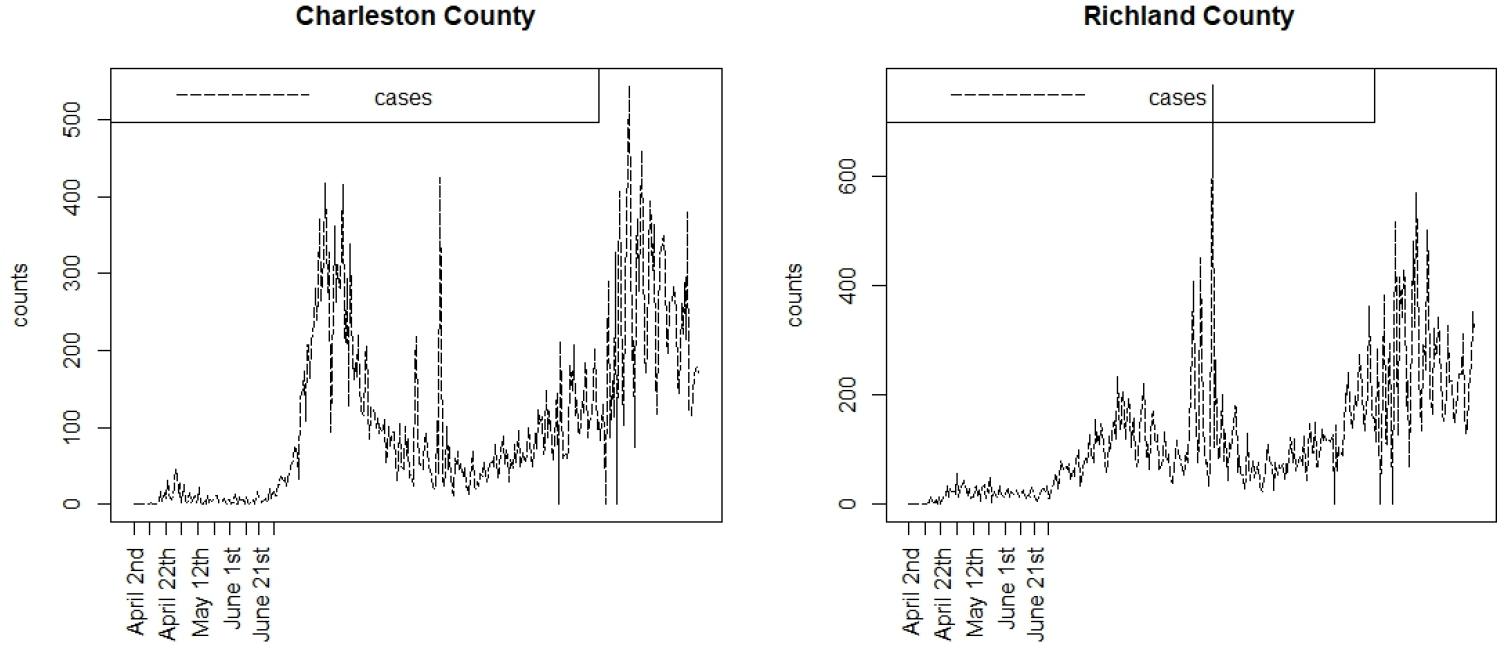
Case count profiles for two South Carolina counties during the first 353 days of the pandemic. The early lockdown dates are shown only.

### Assessing the effects of NP Interventions

It is clear that NPIs have to be compared to situations where interventions have not been introduced. This leads to a difficulty in that finding a suitably matched location or time period with null conditions which can be used as a comparator is crucial. With time series it is possible forecast future outcomes based on currently observed data. As an extension to this it is sometimes useful to make predictions based on lagged observations when current data or recent data is lacking. This prediction is termed nowcasting^1,2^. It has been applied extensively in economic research and is now being adopted in infectious disease epidemiology for making health outcome predictions^3.^. More recently, during the Covid-19 pandemic, the use of nowcasting has been proposed to generate predictions for modifications of social mobility during NPIs^4^. An area which has not been examined is the use of nowcasting to make counterfactual predictions of health outcome events. In particular, the use of observed case count data to predict case counts which are altered by NPIs could be a useful approach in understanding the effect of such interventions.

In this paper we employ nowcasting with Bayesian spatio-temporal models in application to the evaluation of the performance of lockdown NPIs at county level in two contrasting states in the US: South Carolina (SC) and New Jersey (NJ). Our choice of state to examine is based on the contrast between the population structure and political structure of the respective states during the pandemic. SC is a southern state which had a Republican governor and a small mainly rural or semi-rural population (5.2 million), whereas NJ is a northern state with a Democrat governor and a large highly urban population (8.88 million). In each state different PIs were adopted and it is our aim to ascertain how effective these were. Our focus is on the case count data only and we do not examine the mortality counterfactuals although these could also be a focus.

In the next section we outline the models evaluated in this study, the generation of counterfactuals and their comparative evaluation using differential metrics. The data used was made available from the NYT GitHUB repository (https://github.com/nytimes/covid-19-data) which has recorded cumulative case and death counts from state departments of health (cases) and national center for health statistics (NCHS) (deaths) during the course of the pandemic.

The data used here is in the form of daily case and death counts for the period of 353 days from 6^th^ March 2020 to 21^st^ February 2021. Death counts are used only for updating the susceptible population within the case count models, and are not themselves modeled.

### The Bayesian spatiotemporal case model

Lawson and Kim (2021) ^5^ proposed a Bayesian spatio-temporal Covid-19 case count model and was evaluated on the first 88 days of the pandemic in SC. Subsequently this model was extended and updated for the analysis of 353 days^6^. The later analysis of the three waves included a wide range of potential models and modeling strategies. Our models for counterfactuals are based on the retrospective analysis results found.

Define the case count *y*_*ij*_ in the *i* th area and *j* th time period. In our example the areas are counties and time period is days. For SC the number of counties is *m* = 46, and for NJ it is *m* = 22. The total time period is T = 353 days.

As the spread of infection is an important component of infectious disease modeling we assume a susceptible – infected – removed (SIR) model for the process. Essentially,

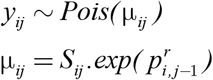

where *S*_*ij*_ is the susceptible population in the *i,j* th unit and 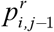 is a propagator which allows transmission as a function of previous counts and related factors.

An example of a simple propagator could be 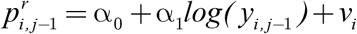 where there is a constant intercept, acting as a log transmission rate, a dependence on the previous infection count in the given county and a final random effect term *v*_*i*_ which allows for extra variation.

Different specifications of 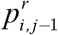 leads to a range of possible models. In these models the susceptible pool evolves over time based on an accounting equation:

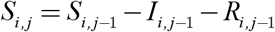

where *I*_*i, j*-1_ is the true infective count at the previous time, which is a function of *y*_*i, j*-1_The relationship between the true infective count and the observed count depends on the level of undetected cases. This could be related to testing frequency and also to unobserved asymptomatic transmission. Previous studies have noted a variety of asymptomatic rates during the pandemic.[e.g. ^7,8^] We assumed a rate of 20% which is a reasonable compromise between the previous levels reported for different population groups. ^8^ Hence we assume that true infective count is a scaled version of observed count: *I*_*i, j*-1_ = λ*y*_*i, j*-1_. The removal term can also be specified as a function of infective numbers. IT is also a function of mortality and so the total removal can specified as *R*_*i, j*_ = γ*I*_*i, j*_ + *d*_*i, j*_ where *d*_*i, j*_ is the current death count. The scaling parameter (γ) can be fixed. In this case it was assumed to be 0.1. However a range of values has been examined for this parameter and the resulting analysis was not affected by this choice.

In previous work^6^, it was found that, out of a range of potential models, for South Carolina counties the model with propagator

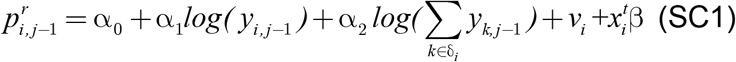

had the lowest WAIC. In this model the 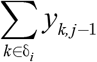 term represents a neighborhood effect (sum of previous count over the neighborhood set δ_*i*_, while 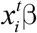 is a linear predictor involving county - level SES predictors (% under the poverty line, % black population, multidimensional deprivation index for 2017 (https://www.census.gov/library/publications/2019/acs/acs-40.html)). In the case of New Jersey, a similar modeling strategy led to the choice of the propagator

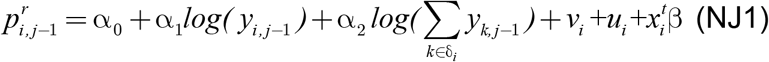

where the term *u*_*i*_ is a spatially correlated effect and 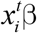 is a linear predictor as above. The spatially correlated term was assumed to follow an ICAR prior distribution^9^, and the uncorrelated effect *v*_*i*_ has a zero mean Gaussian distribution:

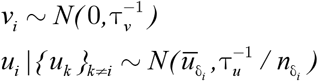

Where 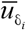 is the mean of *u* in the neighborhood of the *i* th county. The model with this *u*_*i*_ term was not selected in the SC example, which suggest that there is more heterogeneity present in the NJ case.

### Death count modeling

Death counts are also observed, and these are usually related to case numbers either current or lagged. It is unlikely that deaths for Covid19 could arise without there being a case reported (at least in the main epidemic period) and so dependence on lagged case counts is a reasonable assumption. The current death count is defined to be *d*_*i, j*_ and once again we assume a Poisson data model so that 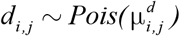. Here the mean death count is parameterized as

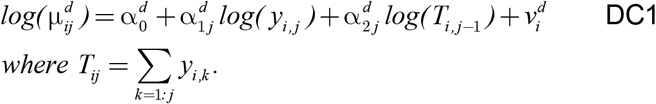

The form of the dependence relies on the need to make the deaths dependent on counts but with a potential lag of undefined length. Hence it is assumed that cumulative case counts should be include as well as the current case number. This model form has been found to provide a good fit to mortality data in the pandemic. ^6,10^

### Nowcasting and Counterfactuals

Nowcasting is often used in situations where infectious disease is being monitored but a reporting delay occurs.^11,12^ This delay can lead to bias such as underreporting or mis-attribution. To alleviate this delay bias, a form of forecasting is used whereby projections of case numbers are made from existing data up to the current time. Once updated data is available, then the count is adjusted. The process is continued until the final time point on study.

This form of missing data forecasting can be applied in other situations. Non-pharmaceutical interventions (NPIs) are often implemented during epidemic periods to try to reduce the spread of disease. These interventions often require spatial restrictions, such as social distancing and mobility constraints such as travel/work bans, or ‘work at home’ mandates and business closures. These are often referred to as lockdowns. During the early part of 2020, many places around the globe implemented lockdowns of various forms to reduce Covid-19 spread. These usually tool the form of gradual business and school closures and final travel bans.

In this paper we examine the use of nowcasting to try to predict the effect of lockdown, or their lifting, on the Covid-19 experience in two contrasting US states: South Carolina (SC) and New Jersey (NJ). SC is a southern state with a small population (∼ 5M) and only small urban centers (Charleston, Columbia, Greenville and Spartanburg). NJ is an urbanized state with a much larger population (∼ 9m) and has (partly) suburban population centers of Trenton, Newark, Jersey City and Atlantic City, bordering the city of New York. We examine the county level case counts of Covid-19 during the lockdown periods relevant to SC and NJ. These periods differ as the state governors decided to implement different types and periods of lockdown. For SC the lockdown started on March 13^th^ and partial lifting of lockdown happened on March 31st (18 days). Final lifting occurred on May 13^th^, but many activities were resumed before this date. For NJ, the lockdown was prolonged until June 9^th^ (80 days), following a partial lockdown from March 9^th^ until March 21^st^. The use of counterfactual generation for Covid-19 NPIs was proposed for employment data previously.^4^ The application of counterfactual generation to spatio-temporal Covid-19 modeling has not been reported before.

### Counterfactual Generation

Consider historical case count data, and assume a good model is known, for these data. We will return to the definition of a good or ‘best’ model at a later stage. For that good model at fixed time (T), a prediction from the model is made. For a spatio-temporal model this prediction is made for all regions under study: in this case counties. Unsupervised prediction for K time units is used to assess what the effect of continuation under a pre T model has compared to the actual observed count over the K time periods. The difference between the observed and counterfactual (predicted) count are then summarized and a comparison is made between SC and NJ state level responses.

The algorithm steps are:

1. Retrospectively fit the ‘best’ model for data up to and including time T.
2. Essentially, we use MCMC sampling from the converged posterior up to T. A large parameter sample is then taken and the SIR count model is allowed to evolve to time T+K, so that a set of predicted counts 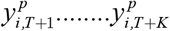 is generated using

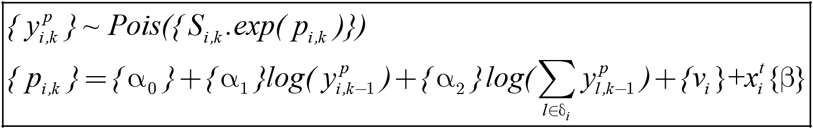

{} denoes the sampled parameter set

δ_*i*_ is the neighborhood set of the *i* th region

This is essentially generating predictions from SC1. For NJ1, an added ICAR term is included. Note that death counts must also be generated, as the case predictions will be a function of the accounting equation which is a function of the concurrent death count. These are generated from the ‘best’ death count model. In this case it is assumed to be DC1.

In this way, a counterfactual is generated in each county and each time period, which can then be compared with the observed count during the NPI. For SC the best model used was that found during a retrospective model search of a wide range of potential models (SC1). A similar search for NJ models led to the use of NJ1 as the ‘best ‘ model’.^6^

### South Carolina counties

We assumed that the crucial time points for this state, measured from the first case, March 6^th^, were T={ 26,42,68}. The first marks the initiation of lockdown, the second the partial lifting and third is the final lifting of lockdown (May13th). Examined were counterfactuals of length 16, 26 and 40, The final end date was June 22^nd^.

Figures 2, 3, 4, and 5 display the results for 4 SC counites at T=26. In these displays the counterfactual is denoted by a thin solid purple line. The 95% credible interval for the counterfactual is shown in purple shading. The mean squared error of the model fit and mean absolute predictive error is also shown. It is notable that for this first period until mid April, Richland is below the observed count and Charleston is mostly higher than observed. Greenville and Spartanburg show a variable picture with many spikes of cases followed by gaps during this period.

**Figure 2.**
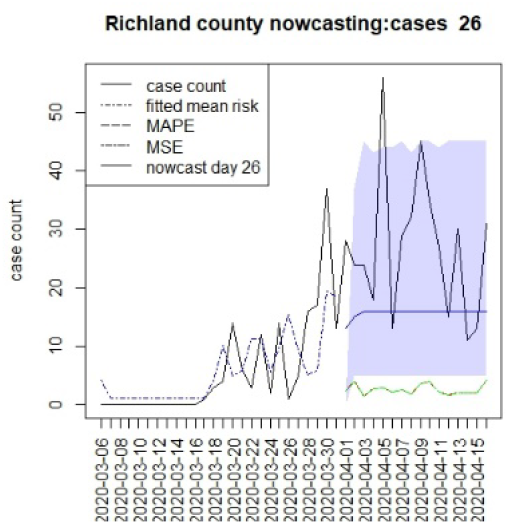
Counterfactual for Richland county at T=26

**Figure 3.**
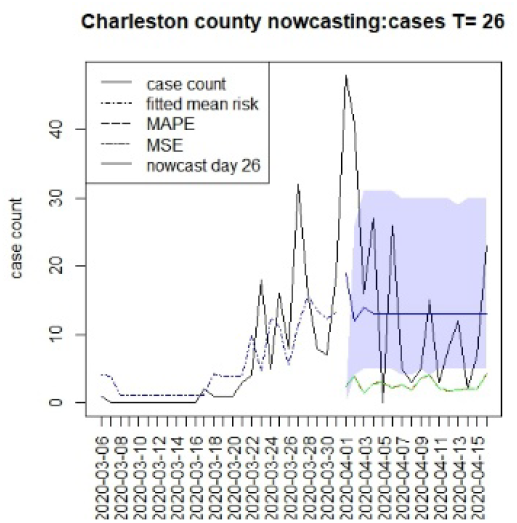
Counterfactual for Charleston county at T=26

**Figure 4.**
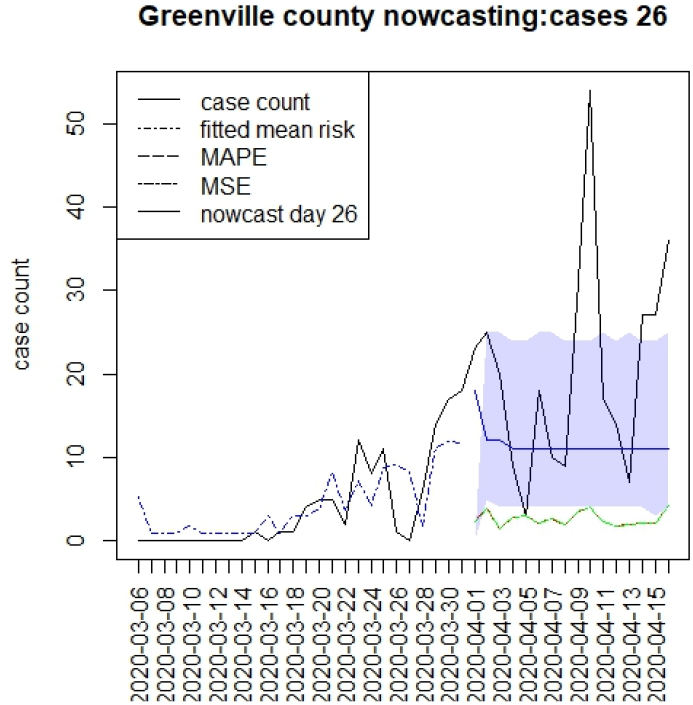
Counterfactual for Greenville county T=26

**Figure 5.**
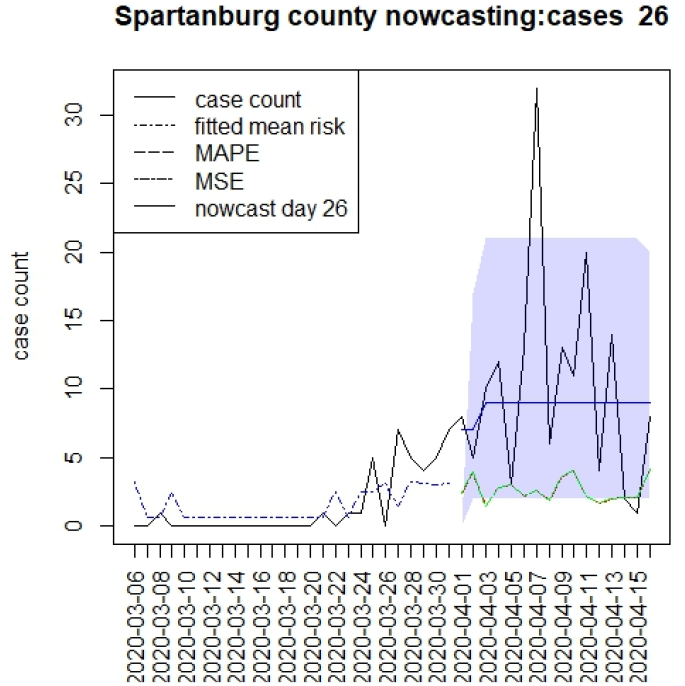
Counterfactual for Spartanburg county at T=26

Figures 6, 7, 8,and 9 displays the counterfactuals for the same four counties at time T=68, which is the end of the lockdown period. We do not display the intermediate case time point here, nor the counterfactuals for deaths, for brevity. While the displays suggest differences between counterfactuals and observed counts, it is more relevant to compute summary measures of the differences. In Table 1 we present results for estimating mean differences between counterfactuals and observed counts. Define the difference at time k as

**Figure 6.**
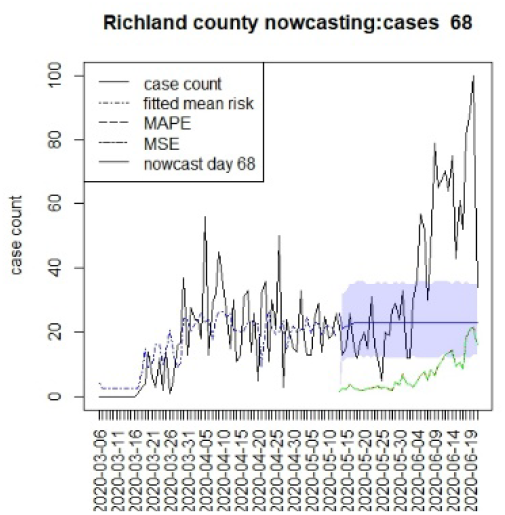
Counterfactual for Richland county T=68

**Figure 7.**
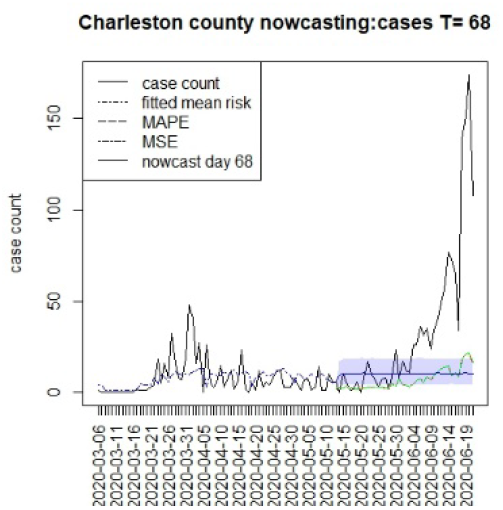
Counterfactual for Charleston county T=68

**Figure 8.**
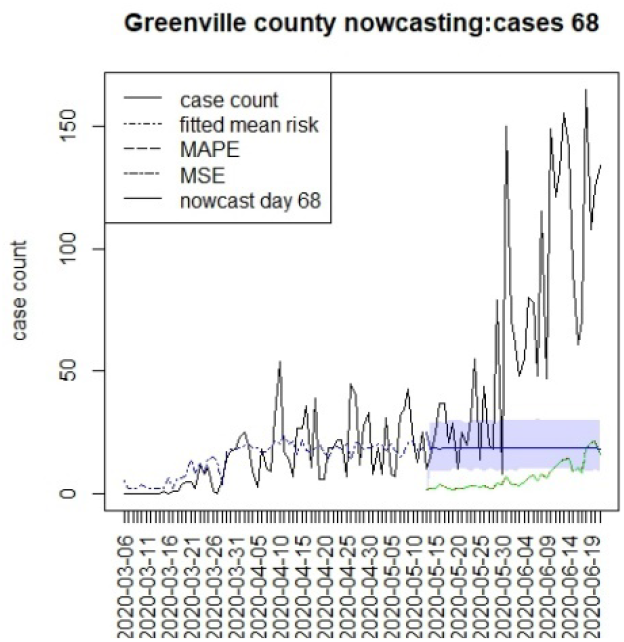
Counterfactual for Greenville county T=68

**Figure 9.**
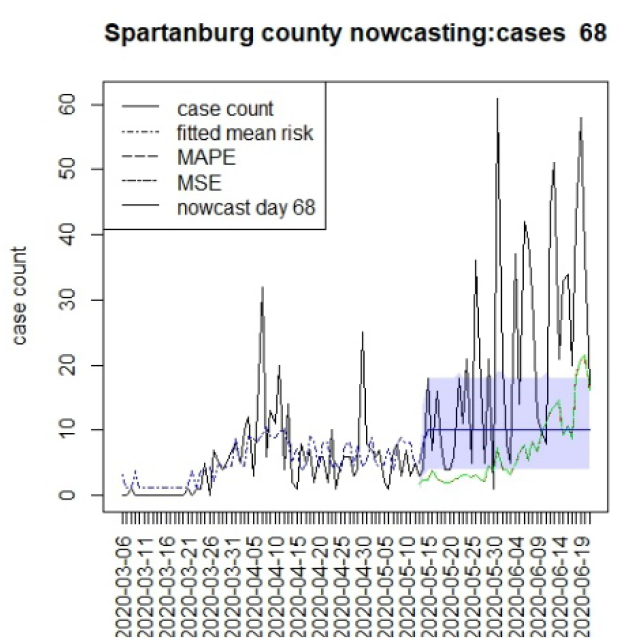
Counterfactual for Spartanburg county T=68

**Figure 9.**
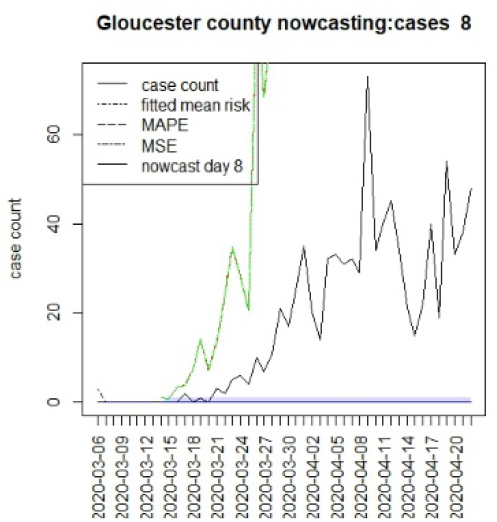
Counterfactual for Gloucester county T=8

**Table 1.** Mean differences between counterfactuals and observed counts averaged over the respective time periods. T is time point, and K is extent. Model assumed is SC1.

| Time | Charleston | Richland | Greenville | Spartanburg |
| --- | --- | --- | --- | --- |
| T26K16 | -1.68 | -11.1 | -9.06 | -1.37 |
| T42K26 | 10.6 | 2.34 | -4.00 | 6.57 |
| T68K40 | -23.8 | -15.3 | -48.6 | -11.4 |

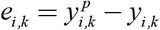 and the mean difference is 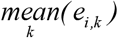

The MAPE is given by *(M)APE*_*i, k*_ = *abs(e*_*i, k*_ ) and the MSE by 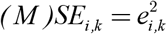.

These time-based loss measures are shown on the counterfactual figures.

In terms of the overall mean levels any negative difference represents a situation where the case load is higher than the predicted counterfactual. This suggests that the in the first period the prediction was everywhere lower than case counts, as there was limited lockdown. In the second period Charleston and Richland achieved positive results as they remained lockdown with lower case numbers, whereas Greenville remained negative. In fact Greenville remained with a high case load throughout out the periods, and this suggest that compliance was poor in this county. Spartanburg had a similar pattern to Richland and Charleston, however. It is important to note that the early lockdowns di not help the case count in the second larger wave during the summer of 2020. All the predictions returned negative mean differences during the final period. It is notable that the predictions across long lags tend to have wide credible intervals and so some degree of uncertainty in these estimates remains. ^13^ In addition, it is also notable that beyond initial step predictions the SIR model leads to almost constant overall risk mean levels. This is typically due to lack of future data support and the need for shocks within a SIR model to allow for peak generation.

### New Jersey counties

We assumed that the crucial time points for this state, measured from March 6^th^, were T={8,16,96}. The first marks the initial restrictions on March 14^th^, and the second the 22^nd^ March when a more restrictive lockdown was imposed. The last time is when the lockdown was finally lifted (June 10^th^). In this case we have examined a 40 day period beyond the T times to examine longer term lockdown effects.

Figures 10 – 21 display the results of fitting the model NJ1 and the posterior expected counterfactuals for the counties of Gloucester, Bergen, Hunterdon and Middlesex.

**Figure 10.**
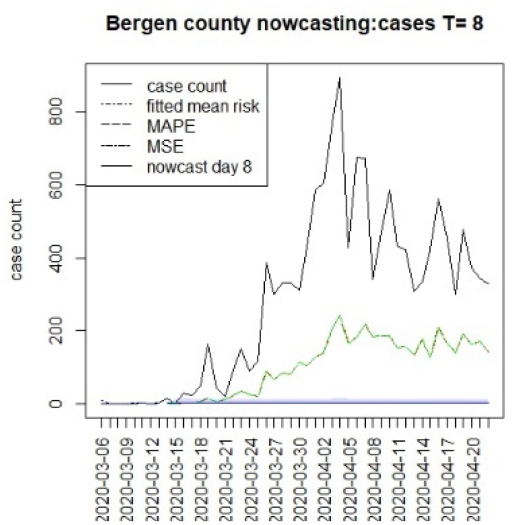
Counterfactual for Bergen county T=8

**Figure 11.**
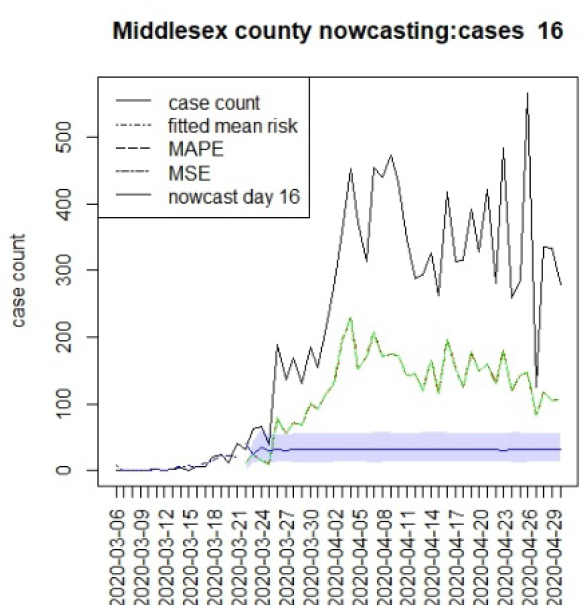
Counterfactual for Middlesex county T=16

**Figure 12.**
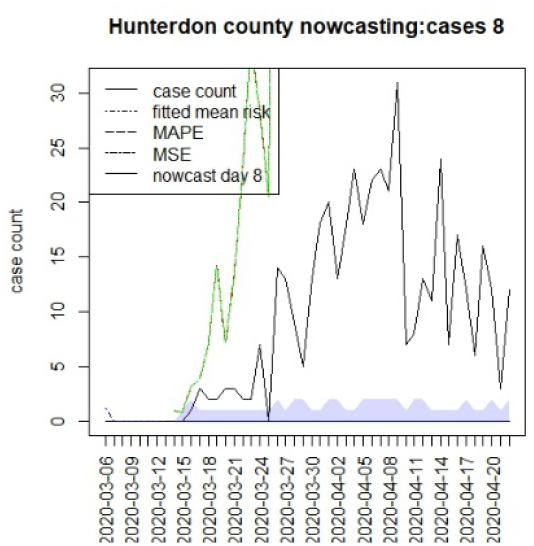
Counterfactual for Hunterdon county T=8

**Figure 12.**
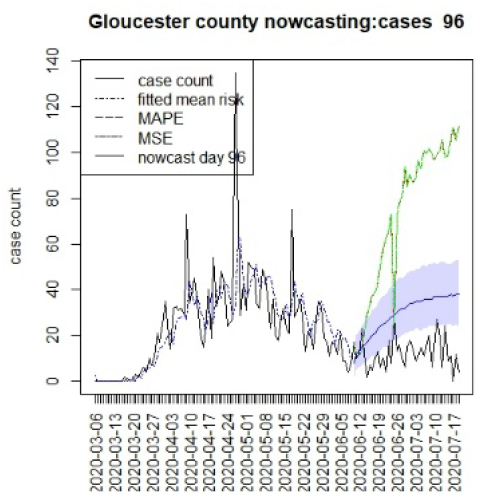
Counterfactual for Gloucester county T=96

**Figure 13.**
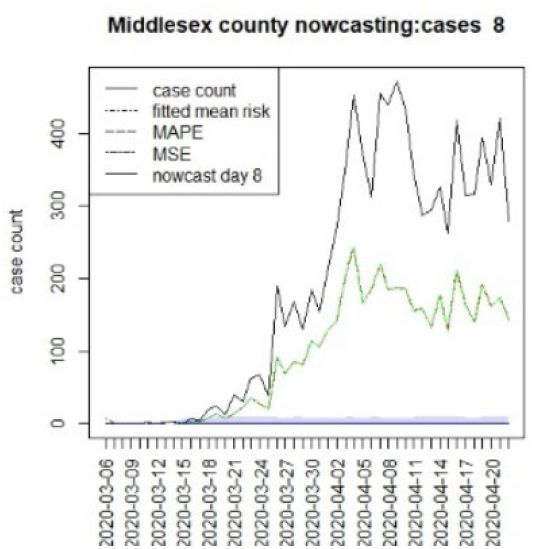
Counterfactual for Middlesex county T=8

**Figure 13.**
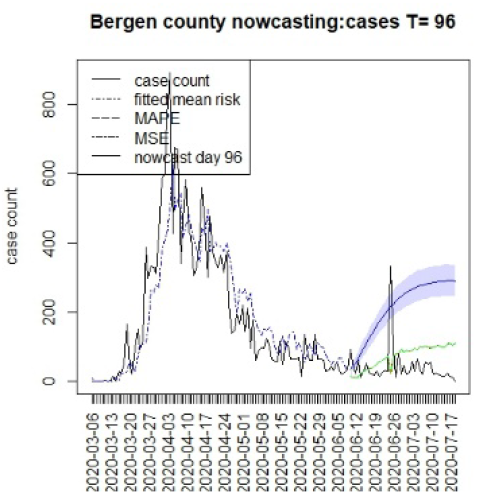
Counterfactual for Bergen county T=96

**Figure 14.**
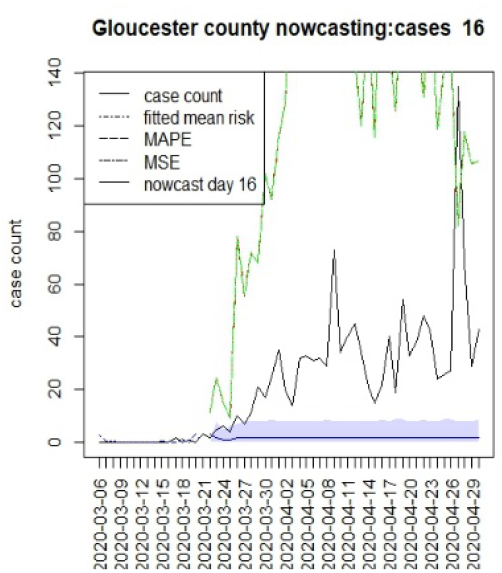
Counterfactual for Gloucester county T=16

**Figure 14.**
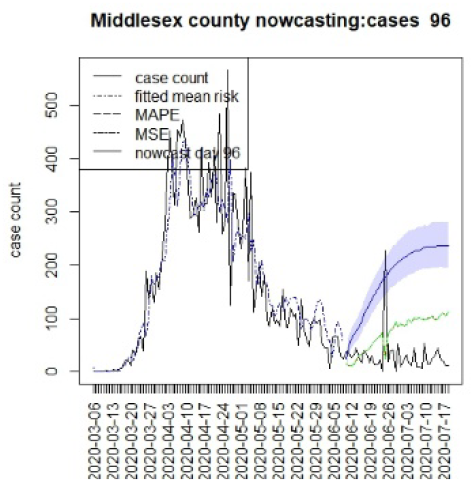
Counterfactual for Middlesex county T=96

**Figure 15.**
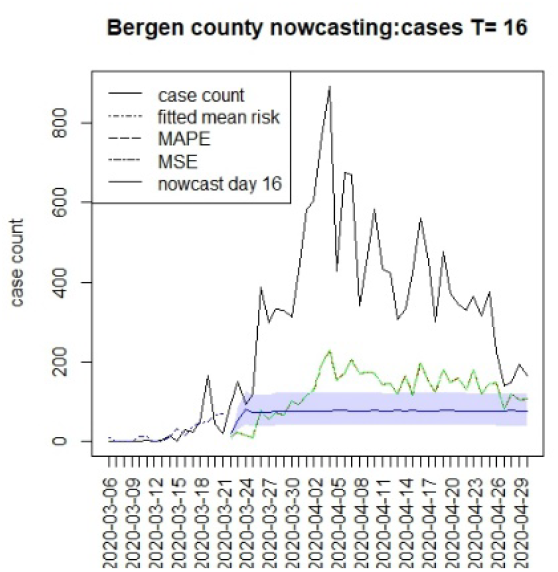
Counterfactual for Bergen county T=16

**Figure 16.**
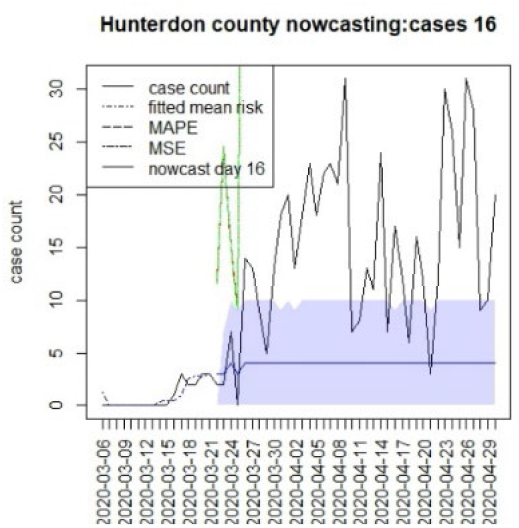
Counterfactual for Hunterdon county T=16

**Figure 20.**
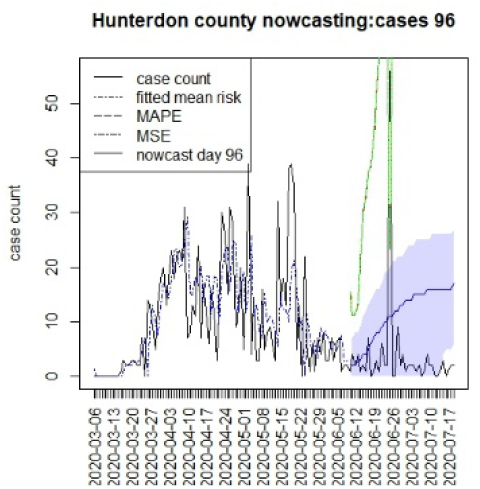
Counterfactual for Hunterdon county T=96

At T=8, it is clear that the nowcasts under report the observed case counts considerably. However at T=16 the situation improved with higher prediction although still mainly below the observed case count. By T=96 the observed cases are below the counterfactual. This is clearly reflected in Table 2 where the differentials become highly positive by T=96. This suggests that at this point the case load has been reduced significantly.

**Table 2.** Mean differences in counterfactuals and observed counts for four NJ counties based on the assumed best model NJ1.

| Time | Bergen | Gloucester | Hunterdon | Middlesex |
| --- | --- | --- | --- | --- |
| T8K40 | -338.6 | -21.4 | -10.9 | -225.1 |
| T16K40 | -306.2 | -29.2 | -10.8 | -258.8 |
| T96K40 | 172.2 | 17.0 | 7.8 | 147.4 |

## Discussion

The counterfactual generation pursued in this paper has a number of drawbacks. First, long term prediction has been shown to demonstrate very wide credible intervals (see e.g. Figures 2-5.) This means that predictions are potentially variable and do not have high confidence. This would appear to be in part because of the SIR model form, but also as data support is limited the further in the future prediction is made. A second issue that arises with SIR model predictions is that the overall risk level becomes relatively constant over time. This is due to the lack of jumps in risk based on the final observed data point. ^13^ Although random effects are commonly used in Bayesian disease mapping as a way to deal with extra variation, there is a trade off as they might not be well estimated when the information fed in the system is too diffused particularly for new emerging diseases. Nonetheless, this work has proposed an extension of SIR Bayesian disease mapping framework to account for uncertainty in infectious surveillance. In addition, the range of prediction should be further examined to find the optimal predictive interval, since this could have an effect on both accuracy and computing resources of surveillance activities in which timeliness is a key.^12^

## Conclusions

The approach proposed here highlights the differentials between both counterfactuals and observed (confirmed) case counts, as well as between regions and states.

With respect to the county differences, there is strong evidence for major differences in response to the interventions between counties in SC. Greenville county in particular shows continual case spread during the lockdowns. In other analysis this the continued existence of clusters of case counts in that county supports the conclusion that non - compliance was common there. The particular difference that is clear appears in the second period after T=26 when Charleston and Richland had reduced case loading whereas Greenville remained above the counterfactual throughout the three periods. In the case of New Jersey, the clear trend was for some success during the middle period and then positive differentials after T=96 across all counties which suggests that suppression was achieved.

While the comparison of states is marred by the fact that the lockdowns were of different kinds and durations. It is quite remarkable that the patterns of compliance are markedly different both between states and within the states. SC did not succeed in locking down adequately and had no NPI in place for the second wave during the summer of 2020. In addition the large difference remained between counties within that state. Whereas, New Jersey maintained their lockdowns and achieved a degree of suppression with similar pattern across counties.

Finally, we note that the approach described here could have sensitivity to choice of T. However, the choice of T is usually defined by policy decisions and so there is only limited possibility to alter these times. Sensitivity to the choice of K could be apparent but we believe that the due to averaging effects across time spans this is limited.

An advantage of this method, is the fact that confidence intervals could be derived for differentials and functions of differentials of various kinds. Here we present basic averages that highlight differences between states and regions. In future work we plan to refine our summarization of the differentials to better reflect the variability. In addition we would examine the use of different data assimilation approaches to improve predictions.

## Data Availability

Data is publically available from GitHub but can also be requested directly from the authors

https://github.com/nytimes/covid-19-data

## References

[1] Johansson MA, Powers AM, Pesik N, Cohen NJ, Staples JE (2014) Nowcasting the Spread of Chikungunya Virus in the Americas. PLOS ONE 9(8): e104915. https://doi.org/10.1371/journal.pone.0104915

[2] Carriero, A., Clark, T. E., & Marcellino, M. (2022). Nowcasting tail risk to economic activity at a weekly frequency. Journal of Applied Econometrics, 37(5), 843– 866. https://doi.org/10.1002/jae.2903

[3] McGough SF, Johansson MA, Lipsitch M, Menzies NA (2020) Nowcasting by Bayesian Smoothing: A flexible, generalizable model for real-time epidemic tracking. PLOS Computational Biology 16(4): e1007735. https://doi.org/10.1371/journal.pcbi.1007735

[4] T.,J., B., N., Middleton, T. (2021) Modeling the Economic and Societal Impact of Non-Pharmaceutical Interventions During the COVID-19 Pandemic. Chance, 34, 2, https://doi.org/10.1080/09332480.2021.1915028

[5] Lawson and Kim (2021) Space-time Covid-19 Bayesian SIR modeling in South Carolina. PlosOne https://doi.org/10.1371/journal.pone.0242777

[6] Lawson and Kim (2022) Bayesian Space-time SIR modeling of Covid-19 in two US states during the 2020-2021 pandemic. PlosOne (accepted)

[7] Sah, P., Fitzpatrick, M., Zimmer, C,. et al (2021) Asymptomatic SARS-CoV-2 infection: A systematic review and meta-analysis. Proceedings of the National Academy of Sciences, 118 (34) e2109229118; DOI: 10.1073/pnas.2109229118

[8] Ma Q, Liu J, Liu Q, et al.(2021) Global Percentage of Asymptomatic SARS-CoV-2 Infections Among the Tested Population and Individuals With Confirmed COVID-19 Diagnosis: A Systematic Review and Meta-analysis. JAMA Netw Open.;4(12):e2137257. doi:10.1001/jamanetworkopen.2021.37257

[9] Lawson, A. B. (2018) Bayesian Bayesian Disease Mapping: hierarchical modeling in spatial epidemiology CRC Press, New York 3rd Ed

[10] Lawson, A. B. (2022) Evaluation of Predictive capability of Bayesian Spatio-temporal models for Covid-19 spread Research Square https://doi.org/10.21203/rs.3.rs-1870683/v1

[11] McGough SF, Johansson MA, Lipsitch M, Menzies NA (2020) Nowcasting by Bayesian Smoothing: A flexible, generalizable model for real-time epidemic tracking. PLOS Computational Biology 16(4): e1007735. https://doi.org/10.1371/journal.pcbi.1007735

[12] Rotejanaprasert, C., Ekapirat, N., Areechokchai, D. et al. Bayesian spatiotemporal modeling with sliding windows to correct reporting delays for real-time dengue surveillance in Thailand. Int J Health Geogr 19, 4 (2020). https://doi.org/10.1186/s12942-020-00199-0

[13] Daza-Torres, M., Capistrán, M., Capella, A., Christen, J., (2022) Bayesian sequential data assimilation for COVID-19 forecasting, Epidemics, 39, 100564 https://doi.org/10.1016/j.epidem.2022.100564.

